# Modelling the role of quarantine escapees on COVID-19 dynamics

**DOI:** 10.1101/2022.07.30.22278240

**Authors:** Josiah Mushanyu, Chinwendu Emilian Madubueze, Williams Chukwu, Zviiteyi Chazuka, Frenick Mudzingwa, Chisara Ogbogbo

**Affiliations:** Department of Mathematics, University of Zimbabwe, Box MP 167, Mount Pleasant, Harare, Zimbabwe; Department of Mathematics/Statistics/Computer Science, University of Agriculture Makurdi, Nigeria; Department of Mathematics and Applied Mathematics, University of Johannesburg, Auckland Park 2006, South Africa; Department of Mathematics, School of Natural Sciences and Mathematics, Chinhoyi University of Technology, Zimbabwe; Department of Mathematics, University of Ghana, Ghana

**Keywords:** COVID-19, Quarantine, Escapees, Numerical simulations

## Abstract

The recent outbreak of the novel coronavirus (COVID-19) pandemic which originated from the Wuhan City of China has devastated many parts of the globe. At present, non-pharmaceutical interventions are the widely available measures being used in combating and controlling this disease. There is great concern over the rampant unaccounted cases of individuals skipping the border during this critical period in time. We develop a deterministic compartmental model to investigate the impact of escapees on the transmission dynamics of COVID-19 in Zimbabwe. A suitable Lyapunov function has been used to show that the disease-free equilibrium is globally asymptotically stable provided ℛ_0_ *<* 1. We performed global sensitivity analysis using the Latin-hyper cube sampling method and partial rank correlation coefficients to determine the most influential model parameters on the short and long term dynamics of the pandemic, so as to minimize uncertainties associated with our variables and parameters. Results confirm that there is a positive correlation between the number of escapees and the reported number of COVID-19 cases. It is shown that escapees are largely responsible for the rapid increase in local transmissions. Also, the results from sensitivity analysis show that an increase in the governmental role actions and a reduction in immigration rate will help to control and contain the disease spread.

## 1. Introduction

The novel Coronavirus (COVID-19) which has taken the world by surprise since its first case in Wuhan China in December 2019 has ravaged Africa and Zimbabwe has felt its portion of the bitter pill. As of the 14th of July 2020, there were 13,141,440 confirmed cases worldwide and 573,349 deaths. The United States of America was the most affected country in the world with 3,481,677 confirmed COVID-19 infections and 138,291 deaths [1]. In Africa, South Africa is currently the epicentre of the coronavirus with 287,796 confirmed cases and 4,172 deaths [2]. Zimbabwe recorded its first COVID-19 case on the 20th of March 2020 and its first death was recorded on the 23rd of March 2020 [3]. Thereafter, the government of Zimbabwe imposed a 21-day nation wide lockdown on the 30th of March 2020 [3]. At the time of drafting this manuscript, Zimbabwe had a record of 1034 confirmed cases with 19 deaths [3].

Recently, Zimbabwe experienced a sharp spike in the number of confirmed COVID-19 cases despite the lockdown. The sharp spike was as a result of the increase in the number of returnees from other countries. As a measure to control the spread of the infection, Zimbabwe closed her borders to normal traffic except that of returnees and essential services. Also, the government imposed a mandatory 21 days quarantine on all returnees, a measure that helped in early identification of quarantined returnees who tested positive. These were immediately placed under mandatory isolation within the country’s isolation healthcare centres. However, some of the returnees escaped from the quarantine centres either after they had tested positive or simply because they did not want to stay at the quarantine centres. This led to a rise in the number of local transmissions. As of 10 July 2020, there were 209 escapees from quarantine centres, the police arrested only 28 of them leaving 181 escapees unaccounted for [4]. Among these escapees it was also reported that some had tested positive implying that there was a high possibility of spreading the infection.

The global economic impact of the novel COVID-19 has brought researchers from across the globe together to fight the virus through various research works. These works include the mathematical modelling of the impact of COVID-19 within countries such as South Africa, China, United States and India just to name a few [5, 6, 7, 8]. Most of the mathematical models created made use of an SEIR modelling framework which only varied in the descriptions of the compartment used and the results obtained from the studies. The main thrust of the mathematical modelling approaches used was to predict the future trend of the COVID-19 virus and provide recommendations to policy makers on the way forward. The role of government action in the control of the spread of COVID-19 within communities was modelled by Mushayabasa *et al*. [5]. In their work, they presented a model that looked at the effects of actions such as imposed social distancing, travel restrictions, quarantine, sanitising and hospitalization on the control of the transmission dynamics of COVID-19. Results from the study indicated that intervention methods by the government have a positive impact on the reduction of infection and the authors also suggested important thresholds by which these intervention strategies must be bound. Nyabadza et al. [9] also modelled the effect of one governmental action (social distancing) on the South African population. Results of their study indicated that if social distancing is relaxed by as much as 2% it could have an impact of 23% rise in infections while an increase in social distance adherence by 2% could effectively reduce infections by 18% [9]. Ambikapathy et al. [10] assessed the effect of the imposed lockdown on the transmission dynamics of COVID-19 in India and results from the model suggested that a 21-day lockdown was effective in the reduction of infections in India and if the government could increase lockdown to 42 days the infections would further be reduced. Other mathematical models of note on the subject matter include [11, 12, 13, 14], just to mention a few.

The model presented in this paper looks at the effects of quarantine measures on the dynamics of infection within the Zimbabwean population. Of particular note, the paper looks at the effects of those who escape from quarantine facilities (herein termed escapees) on the dynamics of COVID-19 in Zimbabwe. To the best of our knowledge, the work presented in this study is novel and of high importance not only to Zimbabwe but to Africa as a whole. The next section presents the model formulation while Section 3 presents model analysis. Numerical simulations and global uncertainty analysis is carried out in Section. The paper is concluded in Section 5.

## 2. Model formulation

The human population is subdivided into 8 distinct classes namely; the susceptible individuals, *S*(*t*), returnees in quarantine who were not exposed to COVID-19, *Q*_1_(*t*), returnees in quarantine who were exposed to COVID-19, *Q*_2_(*t*), locals who are exposed to COVID-19, *E*(*t*), undetected infected individuals, *I*(*t*), detected and isolated individuals, *I*_*D*_(*t*), recovered individuals, *R*(*t*) and deceased individuals, *D*(*t*). We assume that the population mixes homogeneously. The returnees coming into the country are classified as *Q*_1_(*t*) and *Q*_2_(*t*), where *Q*_1_(*t*) denotes returnees who undergo quarantine within the country’s centres as prescribed by the government but are found to be COVID-19 negative after completing quarantine. Upon completion of quarantine, these individuals are assumed to enter into class *S*(*t*) at a rate, *θ*, as they still remain susceptible to local infections. The recruitment rate of returnees is given by Λ. A proportion Λ join the class *Q*_1_(*t*) and the remainder (1 - Π) join the class *Q*_2_(*t*). The model assumes that a proportion *qϕ* of individuals in *Q*_2_(*t*) will escape quarantine while positive and hence join class *I*(*t*) of infected individuals, where *q* is the probability of escape and *ϕ* is the escape rate. The remaining proportion (1 - *q*)*ϕ* from *Q*_2_(*t*) undergo testing and may be found positive and proceed to join class *I*_*D*_(*t*). The rationale here is that returnees are tested: at the beginning of quarantine, during quarantine and towards the end of quarantine. At any time during these prescribed periods, an individual may be found positive and proceed to class *I*_*D*_(*t*) or they can escape while positive avoiding further probation. It is assume that the exposed individuals who are locals, *E*(*t*), become undetected infected individuals at a rate, *σ* because they are yet to detected via testing while individuals infected with COVID-19 in class *I*(*t*) are detected through testing at a rate, *δ*_1_ and isolated. Since the beginning of the outbreak, scientists have suggested the possible mutation of the SARS-Cov-2 virus [15]. According to their research, these mutations may cause the weakening of the strength of the virus. At the same time, some individuals may develop resistance and can fight off the virus without seeking medical attention. We thus assume that the recovery of undetected infected individuals, *I*(*t*), occurs at a rate, *δ*_2_ and move to class *R*(*t*). However, a certain proportion of infected individuals will succumb to the infection at a rate *δ*_3_ and move into class *D*(*t*), which is the death class. A proportion, *d*, of individuals that have been detected (and isolated) in *I*_*D*_(*t*) recover at a rate *ρ* and join class *R*(*t*) of recovered individuals. The remaining proportion (1 - *d*)*ρ* succumb to the infection and join class *D*(*t*). We assume permanent immunity of recovered individuals. The model has two infectious compartments, *I*(*t*) and *I*_*D*_(*t*). Hence, we assume that the force of infection for the model is given by

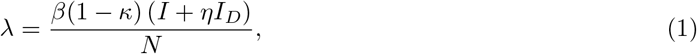

where *β* is the transmission rate of COVID-19, *η* is the modification parameter, where 0 ≤ *η* ≤ 1 and the term (1 - *κ*) represents the effects of governmental action, where 0 ≤ *κ* ≤ 1. A value of *κ* close to 1 indicates that governmental action is effective while a value of *κ* close to zero implies that there is little or no governmental action. The population total is given by

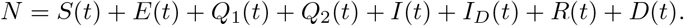

The model dynamics can be summarized as in Figure 1.

**Figure 1:**
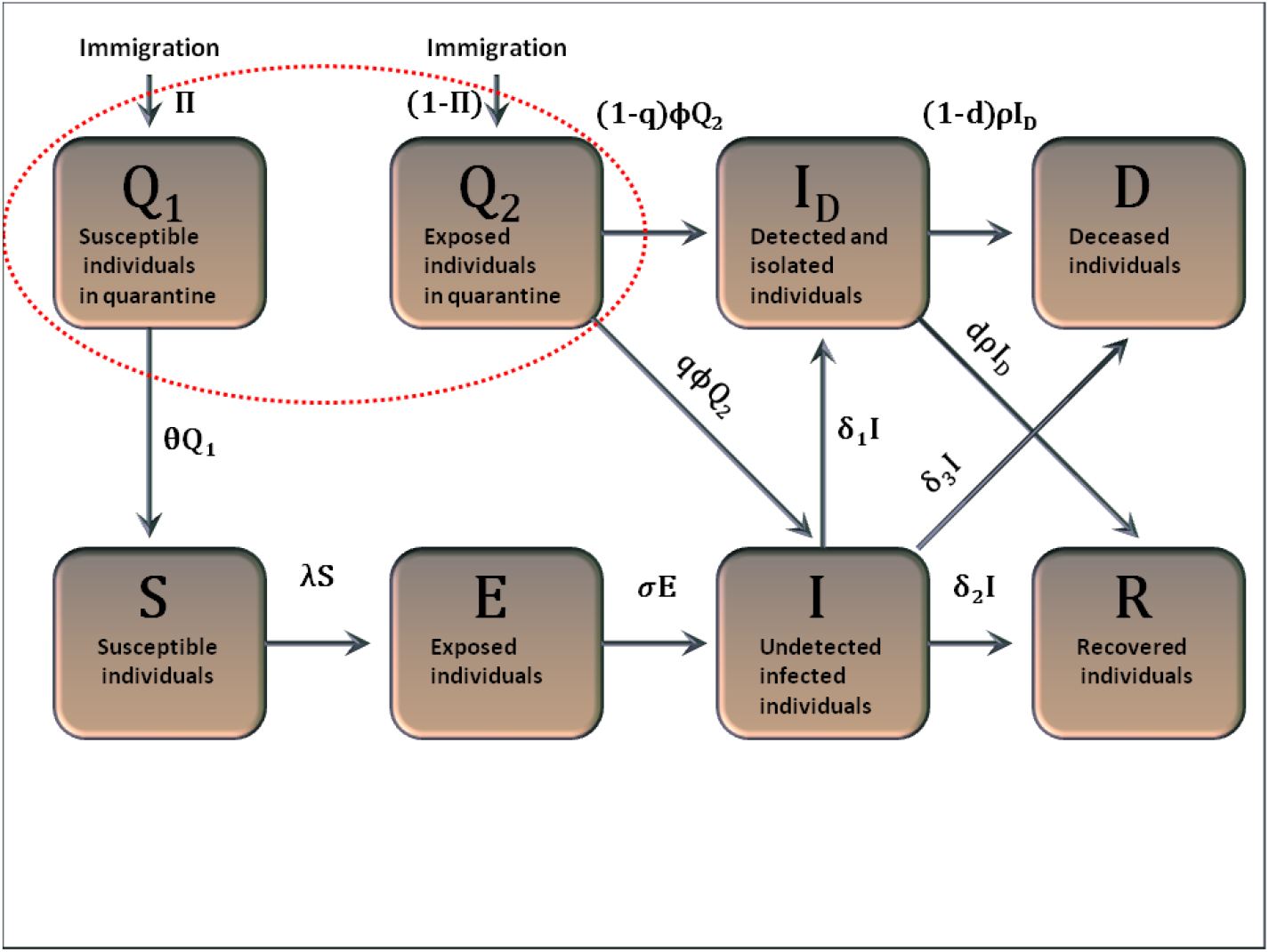
Model flow diagram. The dotted red oval indicates quarantined individuals.

The description of model variables, parameters and assumptions combined with the model flow diagram (Figure 1), leads to the following set of non-linear ordinary differential equations:

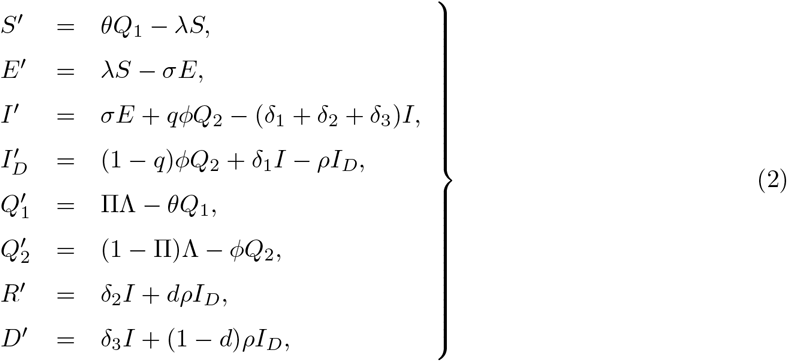

with the initial conditions

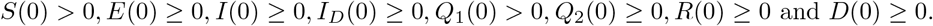

Here, all the model parameters are considered to be non-negative.

Figure 1 correctly captures the epidemiological status of the returnees. However, for simplicity, we assume that all quarantined uninfected individuals will eventually join the class *S*(*t*) of susceptibles as shown in Figure 1. This basically means incorporating in the susceptible population some people who are in quarantine. The error that results from this consideration is negligible. Further, we assume that all the remaining quarantined individuals will either escape to join the class *I*(*t*) or move to the class of detected and isolated individuals upon being screened through testing which is a similar modification done for SARS model by Gumel et al. [16]. This is related to this work in the aspect of being a respiratory disease like COVID-19 and they considered also the impact of undetected entry of infected individuals on the SARS transmission dynamics. Thus, the modified model can be represented by Figure 2 below.

**Figure 2:**
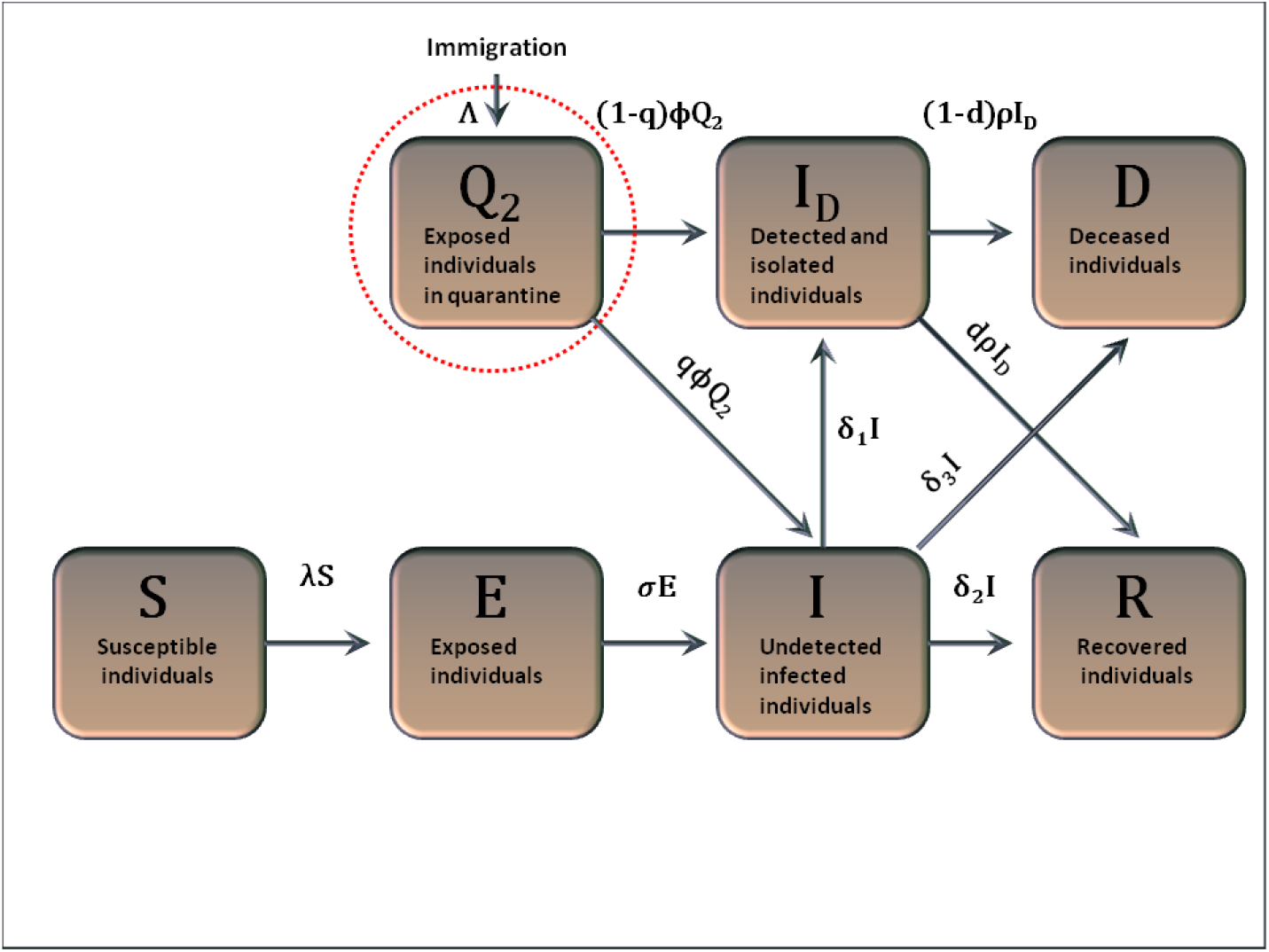
Model flow diagram. The dotted red oval indicates quarantined individuals.

**Figure 3:**
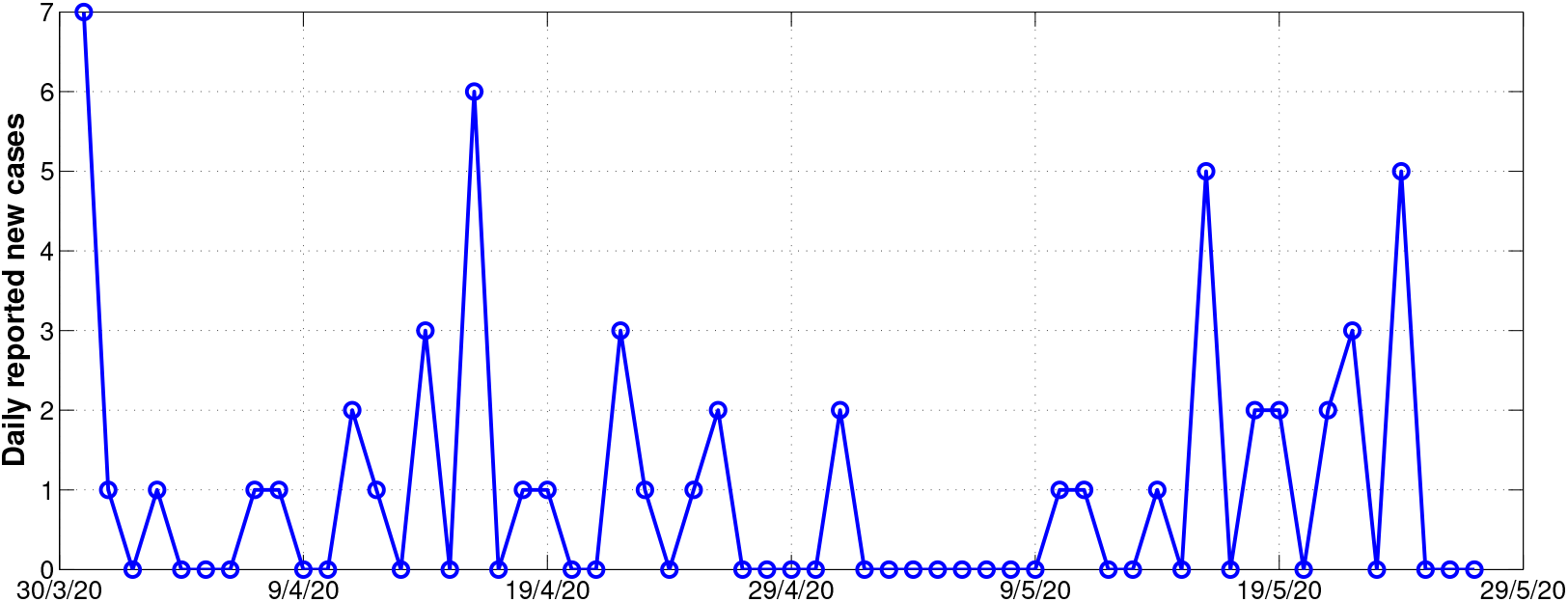
Daily reported new cases in Zimbabwe starting on the 30th of March 2020 and ending on the 27th of May 2020.

**Figure 4:**
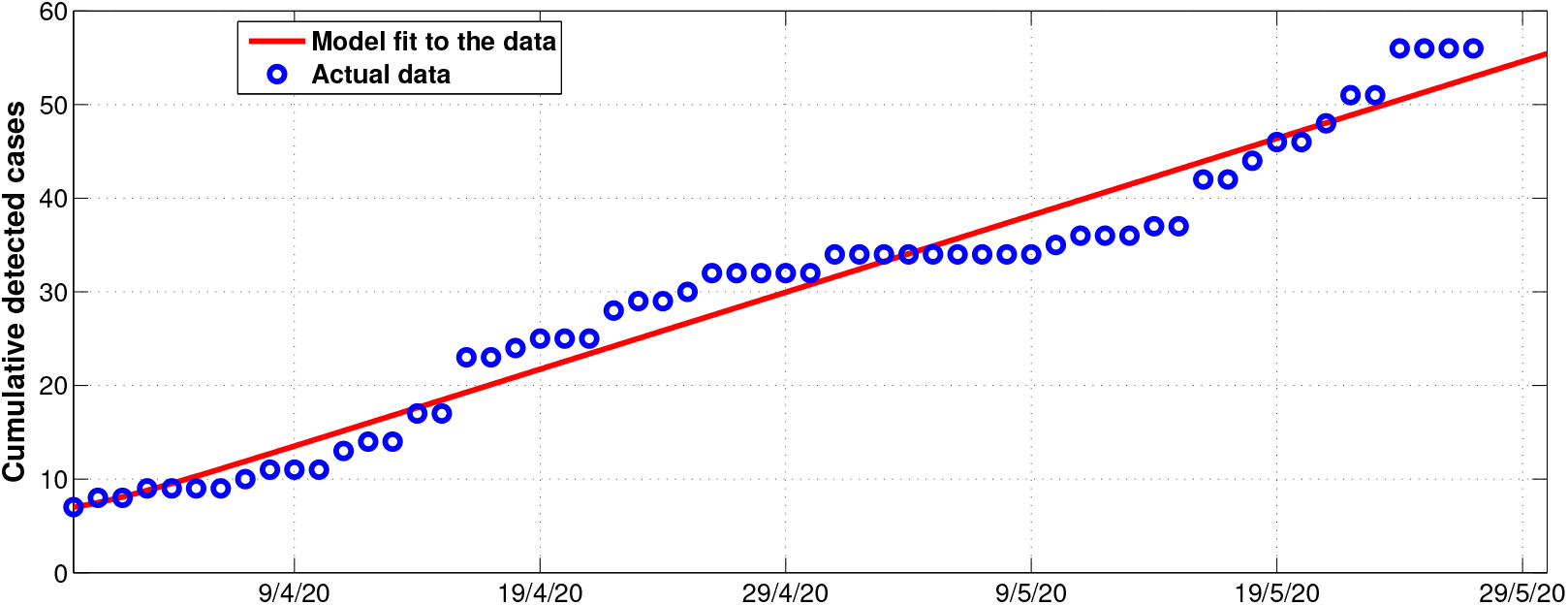
Model system (3) fitted to data for cumulative COVID-19 cases in Zimbabwe before the massive surge of returnees starting from the 30th of March 2020 and ending on the 27th of May 2020. See Table 2. This period covers the whole of the 1st lockdown period, the whole of the 2nd lockdown period and partly covers the initial stages of the 3rd indefinite lockdown period. The *blue circles* indicate the actual data and the *solid red line* indicates the model fit to the data. The initial conditions obtained from data fitting are as follows: *N* (0) = 14 *×* 10^6^; *S*(0) = *N* (0) *- E*(0) *- I*(0) *- I*_*D*_(0) *- Q*_2_(0) *- R*(0); *E*(0) = 0; *I*(0) = 0; *I*_*D*_(0) = 7; *Q*_2_(0) = Λ = 61; *R*(0) = 0; *D*(0) = 1 where *t* = 0 corresponds to the 30th of March 2020 for this case.

**Figure 5:**
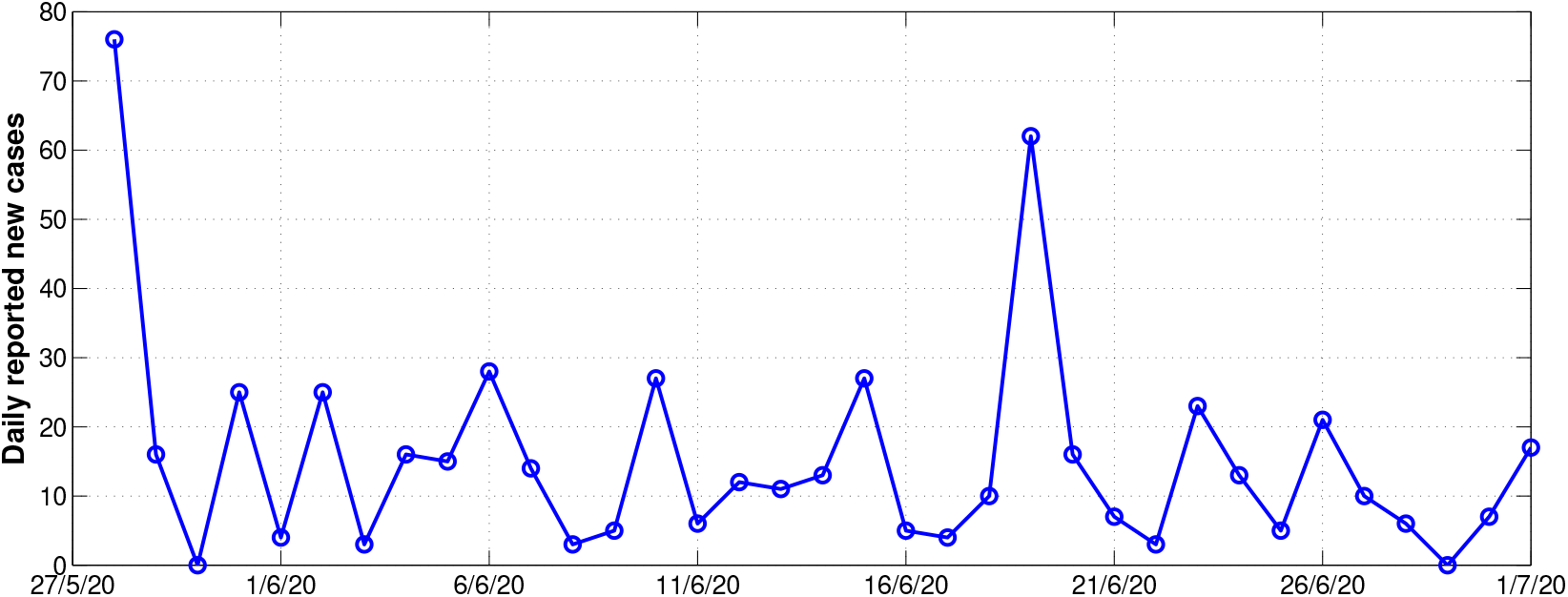
Daily reported new cases in Zimbabwe starting on the 27th of May 2020 and ending on the 30th of June 2020.

**Figure 6:**
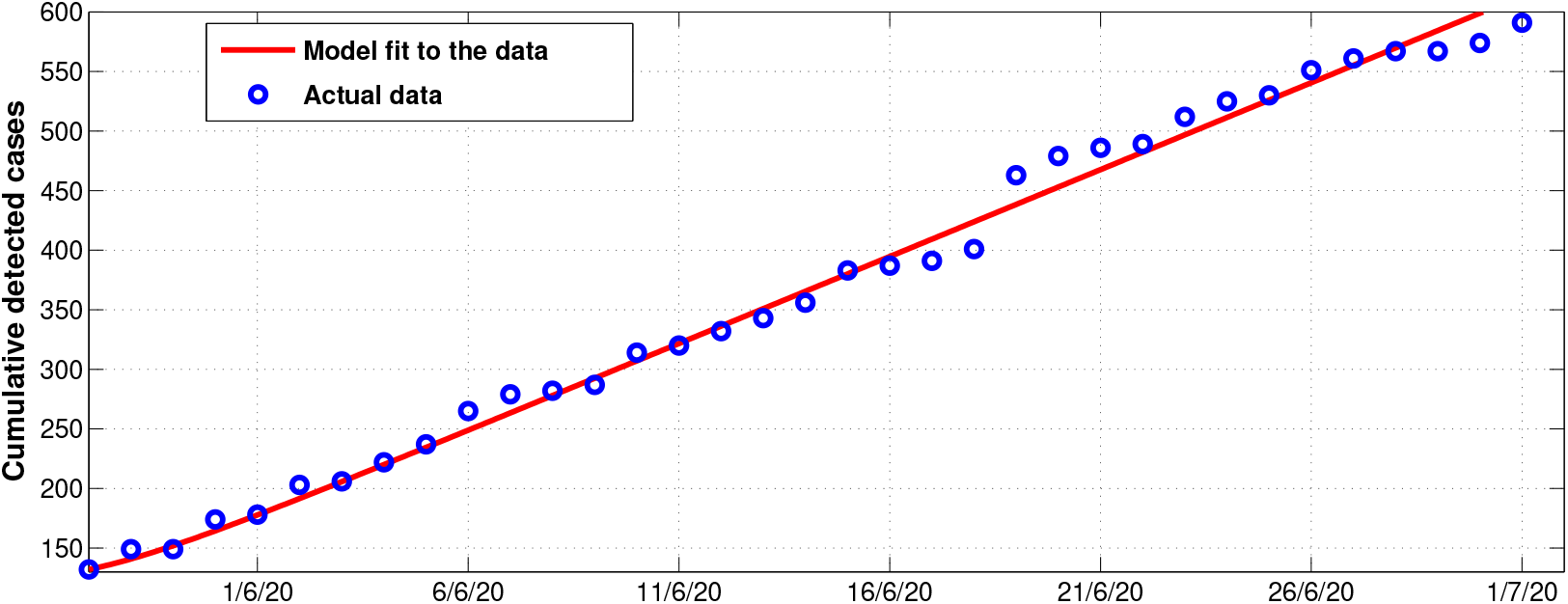
Model system (3) fitted to data for cumulative COVID-19 cases in Zimbabwe after the massive surge of returnees starting from the 27th of May 2020 and ending on the 30th of June 2020 (see Table 3). This period covers the most part of the 3rd indefinite lockdown period. The *blue circles* indicate the actual data and the *solid red line* indicates the model fit to the data. The initial conditions obtained from data fitting are as follows: *N* (0) = 14 *×* 10^6^; *S*(0) = *N* (0) *- E*(0) *- I*(0) *- I*_*D*_(0) *- Q*_2_(0) *- R*(0); *E*(0) = 50; *I*(0) = 0; *I*_*D*_(0) = 132; *Q*_2_(0) = Λ = 1000; *R*(0) = 25; *D*(0) = 4 where *t* = 0 corresponds to the 27th of May 2020 for this case.

We have the following set of nonlinear ordinary differential equations for the modified model:

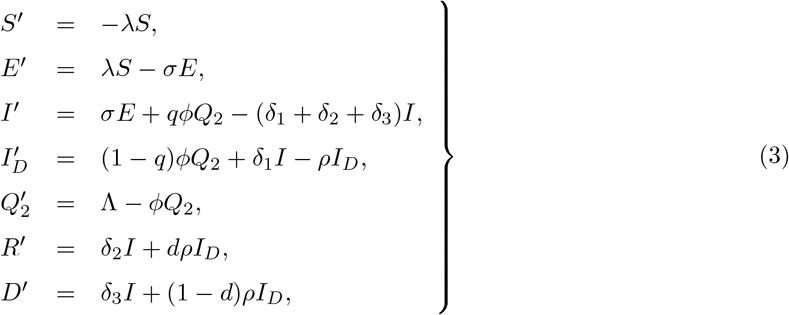

with the initial conditions

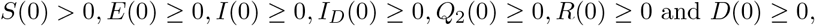

where all the model parameters are considered to be non-negative. Here, Λ is the recruitment of COVID-19 exposed individuals returning back into the country.

## 3. Model analysis

### 3.1. Positivity of solutions

We now consider the positivity of system (3). We show that all the state variables remain non-negative and the solutions of system (3) with positive initial conditions remain positive for all *t >* 0.

#### Theorem 1.

*Given that the initial conditions of system (3) are S*(0) *>* 0, *E*(0) ≥ 0, *I*(0) ≥ 0, *I*_*D*_(0) ≥ 0, *Q*_2_(0) ≥ 0, *R*(0) ≥ 0 *and D*(0) ≥ 0, *then for all t >* 0, *all solutions of model system (3) remain positive in* 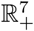.

*Proof*. Following the approach in Gweryina et al. [17], we assume that

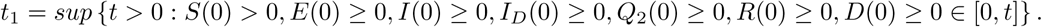

Thus, *t*_1_ *>* 0 and it follows from the first equation of system (3) that

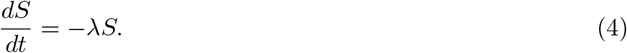

Solving this using separation of variables yields

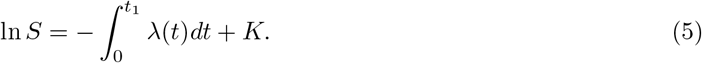

Hence,

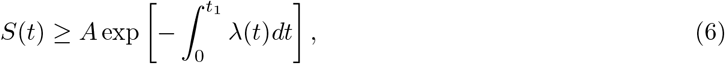

where *A* is a constant to be determined. Therefore, applying initial conditions *S*(0) = *S*_0_ yields *A* = *S*_0_ such that

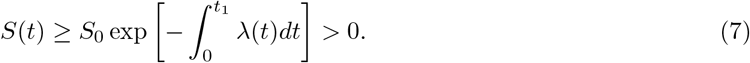

Therefore, it can be shown in a similar manner that *E*(0) ≥ 0, *I*(0) ≥ 0, *I*_*D*_(0) ≥ 0, *Q*_2_(0) ≥ 0, *R*(0) ≥ 0 and *D*(0) ≥ 0, for all *t >* 0. This concludes the proof. □

### 3.2. Invariant region

#### Theorem 2.

The region

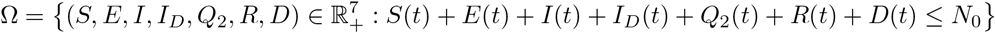

*is positively invariant for system (3), with non-negative initial conditions where N*_0_ *is the initial population*.

*Proof*. Adding all the equations of system (3) yields

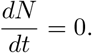

Then, 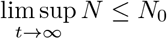. Thus, we have the feasible region for system (3) defined by

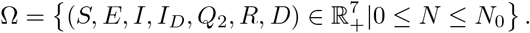

Our analysis will be based on the dynamics in Ω. □

### 3.3. Disease-free equilibrium state and the basic reproduction number

The model has a disease-free equilibrium state

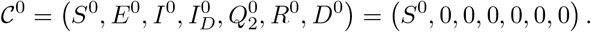

Following the next generation matrix approach by van den Driessche and Watmough [18], we have

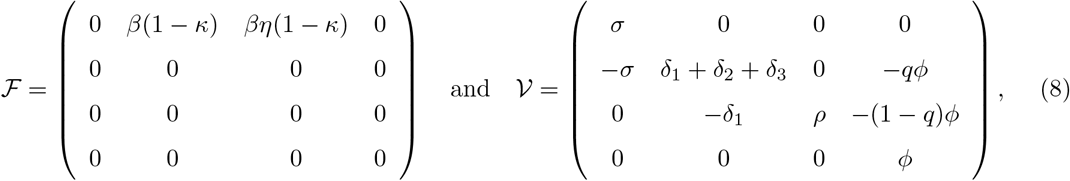

giving

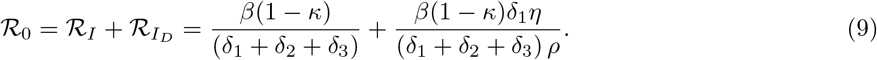

ℛ_*I*_ is the reproduction number contributed by the infected class, *I*(*t*) while 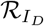 is the reproduction number contributed by the *I*_*D*_ (*t*) class. The term 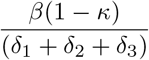 means that the individuals in the undetected infected class *I*(*t*) have contact with the susceptible class at a rate, *β*(1 − *κ*), and they will spend a mean time of 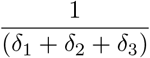 within the undetected Infected class, *I*(*t*). It can be noted that governmental action *κ* will reduce the transmission rate, *β*. The term 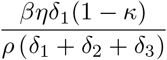 means that some individuals within the undetected Infected class, *I*(*t*) will be detected positive and isolated, at a testing rate of *δ*_1_ and spend a mean time 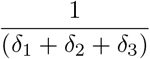 within, *I*(*t*) class. Thereafter, they will progress to the *I*_*D*_(*t*) class and spend an average infectious time of 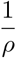 within the *I*_*D*_(*t*) class. The contact rate of such individuals is *βηδ*_1_(1 - *κ*), where *η* is the modification parameter for the *I*_*D*_(*t*) class.

The following result follows from van den Driessche and Watmough [18].

#### Theorem 3.

The disease-free equilibrium for system (3) is locally asymptotically stable provided that ℛ_0_ < 1.

#### 3.3.1. Global stability of the disease-free equilibrium state

We shall now prove the global stability of the disease-free equilibrium point, C^0^ whenever the reproduction number is less than unity.

##### Theorem 4.

The disease-free equilibrium for system (3) is globally asymptotically stable provided that ℛ_0_ < 1.

*Proof*. Using the approach in Shuai and van den Driessche [19], we construct the following Lyapunov function given by

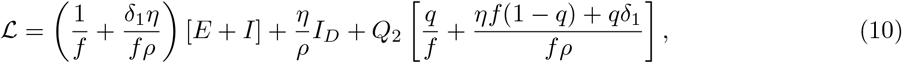

where *f* = *δ*_1_ + *δ*_2_ + *δ*_3_. Differentiating ℒ yields

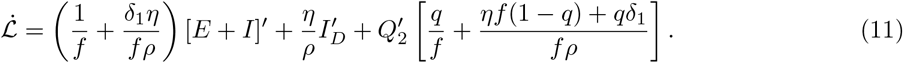

Substitution expressions for 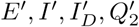 leads to

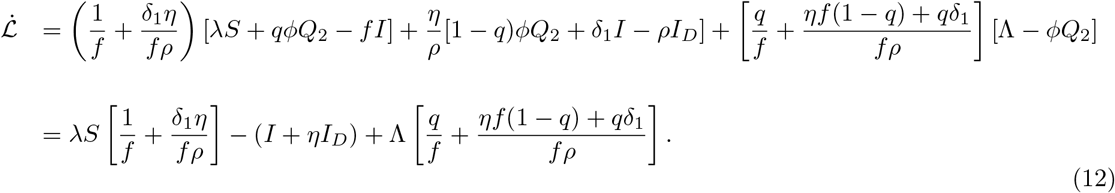

Recall that 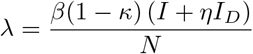 and 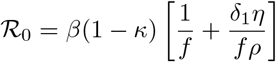. Hence, we obtain

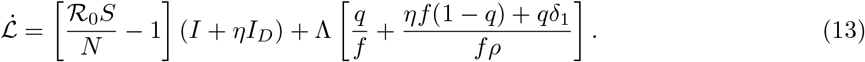

At the disease-free equilibrium Λ = 0 (there are no returnees being recruited into quarantine centres in Zimbabwe), then

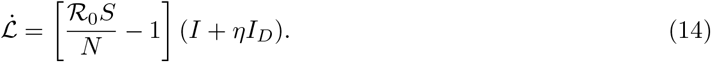

Then, it follows that

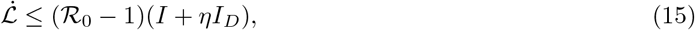

since 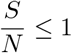. This implies that 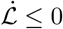 provided that ℛ _0_ ≤ 1 and 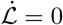 iff *I* = *I*_*D*_ = *E* = *Q*_2_ = 0. Hence, by applying LaSalle’s invariance principle [20] it suffices to conclude that the disease-free equilibrium, 𝒞 ^0^ is globally asymptotically stable provided ℛ_0_ *<* 1 and unstable otherwise. This concludes the proof. □

#### 3.4. Sensitivity analysis of ℛ_0_

We perform sensitivity analysis to establish which parameters have a positive or negative effect on the ℛ_0_ expression given in (9). We use the following normalized forward sensitivity index formula as given in [21]

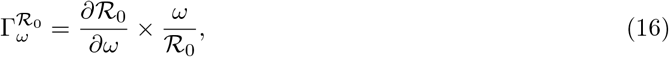

where *ω* is the parameter of interest. Below are sensitivity indices of ℛ_0_ with respect to each of its parameters.

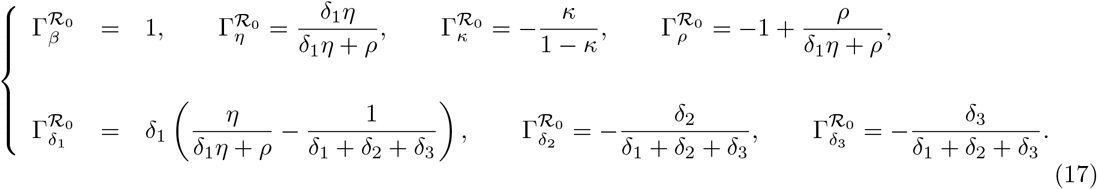

Here, the sensitivity indices for the given parameters as presented in (17) only indicate the direction of influence without quantifying the magnitude of influence of the given parameter. We note that the parameters *β* and *η* have a direct proportional relationship with ℛ_0_, with ℛ_0_ most sensitive to changes in *β*. An increase (or decrease) in either of these parameters will result in an equivalent increase (or decrease) in the value of ℛ_0_. Parameters *κ, ρ, δ*_2_ and *δ*_3_ have an inverse proportional relationship with ℛ_0_, an increase in the values of these parameters will lead to a decrease in the value of ℛ_0_. However, it is unethical to put down measures that will increase the value of *δ*_3_. Thus, measures such as increased surveillance and isolation of infected individuals (increase in *ρ*), and enforcing governmental actions such as wearing of face masks, use of sanitisers, social distancing etc (increase in *κ*) will be of great help in curtailing the spread of COVID-19. Interestingly, 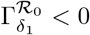 when *ρ >* (2*δ*_1_ + *δ*_2_ + *δ*_3_) *η* and 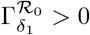 when *ρ <* (2*δ*_1_ + *δ*_2_ + *δ*_3_) *η*.

## 4. Numerical simulations

In this section, we perform numerical simulations of system (3). We consider a case study of COVID-19 transmission dynamics in Zimbabwe. The model considered in the present study has many parameters whose values need to be ascertained in order to properly capture the current COVID-19 transmission dynamics. In order to capture the current COVID-19 dynamics in Zimbabwe, we fit the model to the currently observed COVID-19 data. We assign reasonable ranges from which parameter values are chosen and we obtain parameter values that give the best fit. These parameters are used to perform our numerical simulations. We also carry out some forecasting of the disease dynamics under certain conditions within the framework of the objectives of the study. For instance, we investigate the long term impact of governmental actions such as lockdown, social distancing, closure of schools and universities etc.

We consider COVID-19 data starting from the 30th of March 2020 (day 1) when the first lockdown was instituted in Zimbabwe up to the 30th of June 2020 (day 93). Important time-lines for the management and control of COVID-19 in Zimbabwe as given by the government authorities are as follows:

1. declaration of state of disaster on the 17th of March 2020 [22],
2. 1st lockdown instituted on the 30th of March 2020 [22],
3. 2nd lockdown instituted on the 6th of May 2020 [23],
4. 3rd lockdown instituted on the 20th of May 2020 for an indefinite period covering the drafting of this manuscript [24].

We tabulate the data for COVID-19 cases in three parts namely: (i) the period covering the 1st lockdown (see Table 1), (ii) the period before the massive surge of returnees (see Table 2. This period covers all of the 1st lockdown period, all of the 2nd lockdown period and part of the initial stages of the 3rd indefinite lockdown period), and (iii) the period after the massive surge of returnees (see Table 3. This period covers the most part of the 3rd indefinite lockdown period).

**Table 1:** Confirmed COVID-19 cases in Zimbabwe before lockdown [25].

| Date | 20/3/20 | 21/3/20 | 22/3/20 | 23/3/20 | 24/3/20 | 25/3/20 | 26/3/20 | 27/3/20 |
| --- | --- | --- | --- | --- | --- | --- | --- | --- |
| Total cases | 1 | 3 | 3 | 3 | 3 | 3 | 3 | 5 |
| Date | 28/3/20 | 29/3/20 |  |  |  |  |  |  |
| Total cases | 7 | 7 |  |  |  |  |  |  |

**Table 2:** Confirmed COVID-19 cases in Zimbabwe before massive surge of returnees. This period covers the whole of the 1st lockdown period, the whole of the 2nd lockdown period and partly covers the initial stages of the 3rd indefinite lockdown period [25].

|  |  |  |  |  |  |  |  |  |
| --- | --- | --- | --- | --- | --- | --- | --- | --- |
| **Date** | 30/3/20 | 31/3/20 | 1/4/20 | 2/4/20 | 3/4/20 | 4/4/20 | 5/4/20 | 6/4/20 |
| **Total cases** | 7 | 8 | 8 | 9 | 9 | 9 | 9 | 10 |
| **Date** | 7/4/20 | 8/4/20 | 9/4/20 | 10/4/20 | 11/4/20 | 12/4/20 | 13/4/20 | 14/4/20 |
| **Total cases** | 11 | 11 | 11 | 13 | 14 | 14 | 17 | 17 |
| **Date** | 15/4/20 | 16/4/20 | 17/4/20 | 18/4/20 | 19/4/20 | 20/4/20 | 21/4/20 | 22/4/20 |
| **Total cases** | 23 | 23 | 24 | 25 | 25 | 25 | 28 | 29 |
| **Date** | 23/4/20 | 24/4/20 | 25/4/20 | 26/4/20 | 27/4/20 | 28/4/20 | 29/4/20 | 30/4/20 |
| **Total cases** | 29 | 30 | 32 | 32 | 32 | 32 | 32 | 34 |
| **Date** | 1/5/20 | 2/5/20 | 3/5/20 | 4/5/20 | 5/5/20 | 6/5/20 | 7/5/20 | 8/5/20 |
| **Total cases** | 34 | 34 | 34 | 34 | 34 | 34 | 34 | 34 |
| **Date** | 9/5/20 | 10/5/20 | 11/5/20 | 12/5/20 | 13/5/20 | 14/5/20 | 15/5/20 | 16/5/20 |
| **Total cases** | 35 | 36 | 36 | 36 | 37 | 37 | 42 | 42 |
| **Date** | 17/5/20 | 18/5/20 | 19/5/20 | 20/5/20 | 21/5/20 | 22/5/20 | 23/5/20 | 24/5/20 |
| **Total cases** | 44 | 46 | 46 | 48 | 51 | 51 | 56 | 56 |
| **Date** | 25/5/20 | 26/5/20 |  |  |  |  |  |  |
| **Total cases** | 56 | 56 |  |  |  |  |  |  |

**Table 3:** Confirmed COVID-19 cases in Zimbabwe after massive surge of returnees. This period covers the most part of the 3rd indefinite lockdown period [25].

|  |  |  |  |  |  |  |  |  |
| --- | --- | --- | --- | --- | --- | --- | --- | --- |
| **Date** | 27/5/20 | 28/5/20 | 29/5/20 | 30/5/20 | 31/5/20 | 1/6/20 | 2/6/20 | 3/6/20 |
| **Total cases** | 132 | 149 | 149 | 174 | 178 | 203 | 206 | 222 |
| **Date** | 4/6/20 | 5/6/20 | 6/6/20 | 7/6/20 | 8/6/20 | 9/6/20 | 10/6/20 | 11/6/20 |
| **Total cases** | 237 | 265 | 279 | 282 | 287 | 314 | 320 | 332 |
| **Date** | 12/6/20 | 13/6/20 | 14/6/20 | 15/6/20 | 16/6/20 | 17/6/20 | 18/6/20 | 19/6/20 |
| **Total cases** | 343 | 356 | 383 | 387 | 391 | 401 | 463 | 479 |
| **Date** | 20/6/20 | 21/6/20 | 22/6/20 | 23/6/20 | 24/6/20 | 25/6/20 | 26/6/20 | 27/6/20 |
| **Total cases** | 486 | 489 | 512 | 525 | 530 | 551 | 561 | 567 |
| **Date** | 28/6/20 | 29/6/20 | 30/6/20 |  |  |  |  |  |
| **Total cases** | 567 | 574 | 591 |  |  |  |  |  |

**Table 4:** Description of parameters in system (3) using data for Table 2.

| Description | Symbol | Range | Baseline value | Source |
| --- | --- | --- | --- | --- |
| Disease transmission rate | $\beta$ | $[0.4, 1]$ | 0.5643 | Data fit |
| Governmental action | $\kappa$ | $[0.4239, 0.8478]$ | 0.55 | [29, 5] |
| Modification factor | $\eta$ | $[0.4, 1]$ | 0.7015 | Data fit |
| Mean incubation period | $\sigma^{-1}$ | 5 – 6 days | 5 | [30, 31] |
| Mean duration in quarantine | $\phi^{-1}$ | 14 days | 14 | [28] |
| Proportion of escapees | $q$ | $[0.00138, 0.025]$ day$^{-1}$ | 0.00138 | [26, 27] |
| Detection rate of infected individuals | $\delta_1$ | $[0, 0.7]$ day$^{-1}$ | $4.0861 \times 10^{-7}$ | Data fit |
| Recovery rate of undetected individuals | $\delta_2$ | $[0.0752, 0.1370]$ day$^{-1}$ | 0.0795 | [32, 33] |
| Disease related death for infected individuals | $\delta_3$ | $[0.03, 0.04]$ day$^{-1}$ | 0.04 | [34] |
| Mean duration in isolation | $\rho^{-1}$ | $[0.03, 0.2]$ days | 0.1 | Data fit |
| Proportion of isolated individuals who recovers | $d$ | $[0.26, 0.5]$ day$^{-1}$ | 0.26 | [26, 27] |
| Immigration of exposed individuals | $\Lambda$ | $[50, 100]$ day$^{-1}$ | 61 | Data fit |

In order to estimate our unknown model parameters and unpack the underlying dynamics of the COVID- 19 pandemic in Zimbabwe, we make use of the curve fitting process to approximately quantify the trend of the outcomes of this pandemic. Here, we fit the equations of approximating curves to the available raw field data on COVID-19 dynamics in Zimbabwe. Generally, it will be observed that the fitting curves for any given data set are not unique. Thus, a curve with minimum possible deviation from all the data points will be desired. The least squares curve fit routine (lsqcurvefit) in Matlab with optimization is used to estimate our unknown model parameters. The procedure requires that a lower and an upper bound be set from which estimates of the unknown parameter values are obtained. Thus, using the available data on cumulative cases for COVID-19 in Zimbabwe over a defined time frame, *t*_*i-*1_ ≤ *t* ≤ *t*_*i*_ (where *t*_*i-*1_ and *t*_*i*_ indicate the beginning and end of the time interval, respectively), we estimate using the function

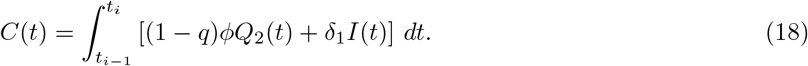

### 4.1. Epidemiological parameter estimation

Under this section, we ascertain epidemiological parameter values used for conducting numerical simulations. We make use of the available expanding literature on COVID-19 dynamics and the Zimbabwe COVID-19 data fitting. Immigration of exposed individuals (Λ) is estimated for the time periods before and after the massive surge of returnees. The mandatory quarantine period for returnees has been set to be 21 days with some individuals escaping from quarantine before this period lapses. Thus, we choose a range of between 14 to 21 days and set the mean duration of quarantine to be *ϕ*^*-*1^ = 14 days [26]. The proportion, *q*, of those escaping from quarantine facilities is chosen to fall within the range [0.00138, 0.025] per day [26, 27]. The proportion of the detected and isolated individuals who progress into the recovered class *R*(*t*) is considered to be within the range *d* = [0.26, 0.5], drawn from the work done in [26, 28]. Following [29, 5], we assign the range for the governmental action parameter *κ* as *κ* = [0.4239, 0.8478]. Further, the remaining parameter values are estimated from the COVID-19 data fit, which are; the disease transmission rate; *β* = 0.5643, modification parameter; *η* = 0.7015, detection rate of infected individuals; *δ*_1_ = 4.0816 × 10^*-*7^ and the mean duration period for isolation of detected individuals is *ρ*^*-*1^ = 10 days. We provide a summary of the description of parameters, ranges and values used in Table 4 below.

### 4.2. Sensitivity analysis

According to Marino [35], we note that in any mathematical modelling exercise, the model variables and parameters are uncertain which makes it difficult to quantify. Also, sensitivity analysis can be carried out over time interval with no particular time frame under investigation for exploratory purposes. For instance, the exact number of exposed returnees remains difficult to accurately measure considering some unaccounted cases of individuals who skip the border. More generally, describing a phenomena via use of models is often linked with incomplete available knowledge due to lack of analogous experimental measures. Hence, we chose only the most important parameters and/or state variables in relation to the subject/aim of study. It is important to note that as indicated in [35], all parameters that have PRCCs between -0.2 to 0.2 are considered to be less sensitive parameters, though this can be further justified by the calculation of the associated P-values in further study. Sensitivity analyses were done using the state variables; the undetected individuals *I*, detected and isolated individuals *I*_*D*_ and the exposed individuals in quarantine *Q*_2_ to determine the most influential parameters; Λ, *ρ, δ*_1_, *q, ϕ, κ* and *σ* on the increase/decrease of the escapee individuals on COVID-19 disease transmission dynamics. We aim to find out the impact of these parameters against the state variable of choice for their short and long term transmission dynamics for our model by considering their Partial Rank Correlation Coefficients (PRCC) values over the modelling time. This will help us to quantify the role of escapees on SARS-CoV-2 transmissions before and after surge for the disease. Here, we consider the sensitivity analysis of these parameters before and after surge of COVID-19 as presented in subsections below.

#### 4.2.1. Sensitivity analysis before surge

Some parameters from Table 1 are used to carry out global uncertainty sensitivity analysis of model system (3) with the combination of Latin Hypercube Sampling (LHS) and the Partial Rank Correlation Coefficients (PRCC) as in [35]. A sample size of *N* = 500 with a unit step of 1 is used to implement the simulations in this subsection for both the short term and long term dynamics of the disease and their results are depicted in Figures 7 and 8 respectively. The basic reproduction number for this case was found to be ℛ_0_ = 2.1.

**Figure 7:**
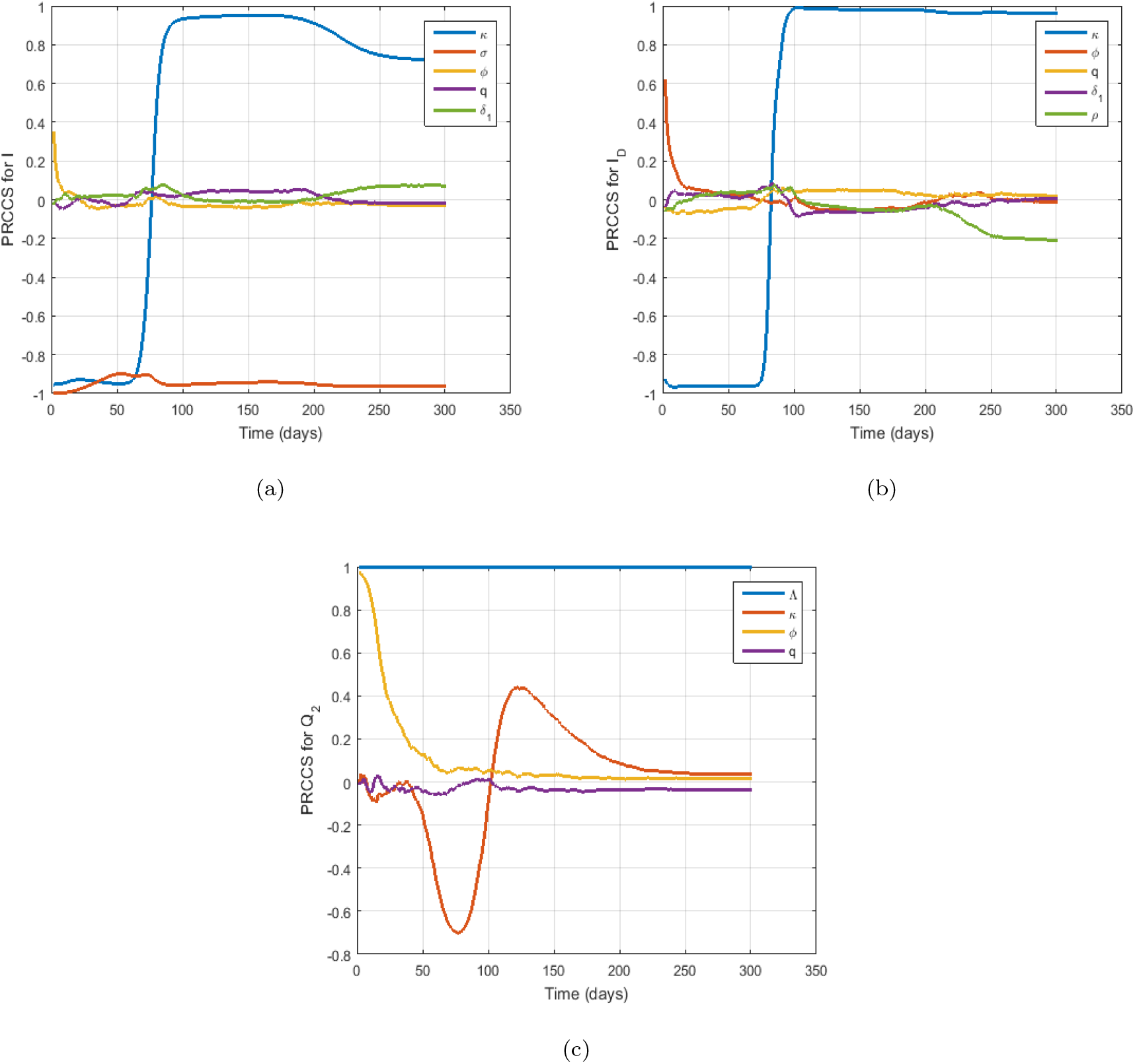
Plots of PRCC values showing the effects of long term dynamics before surge for parameter values on COVID-19 disease in Zimbabwe over time for (a) parameters *κ, σ, ϕ, q, δ*_1_ against *I*. (b) parameters *κ, ϕ, q, δ*_1_, *ρ* against *I*_*D*_. (c) parameters Λ, *κ, ϕ, q* against *Q*_2_.

**Figure 8:**
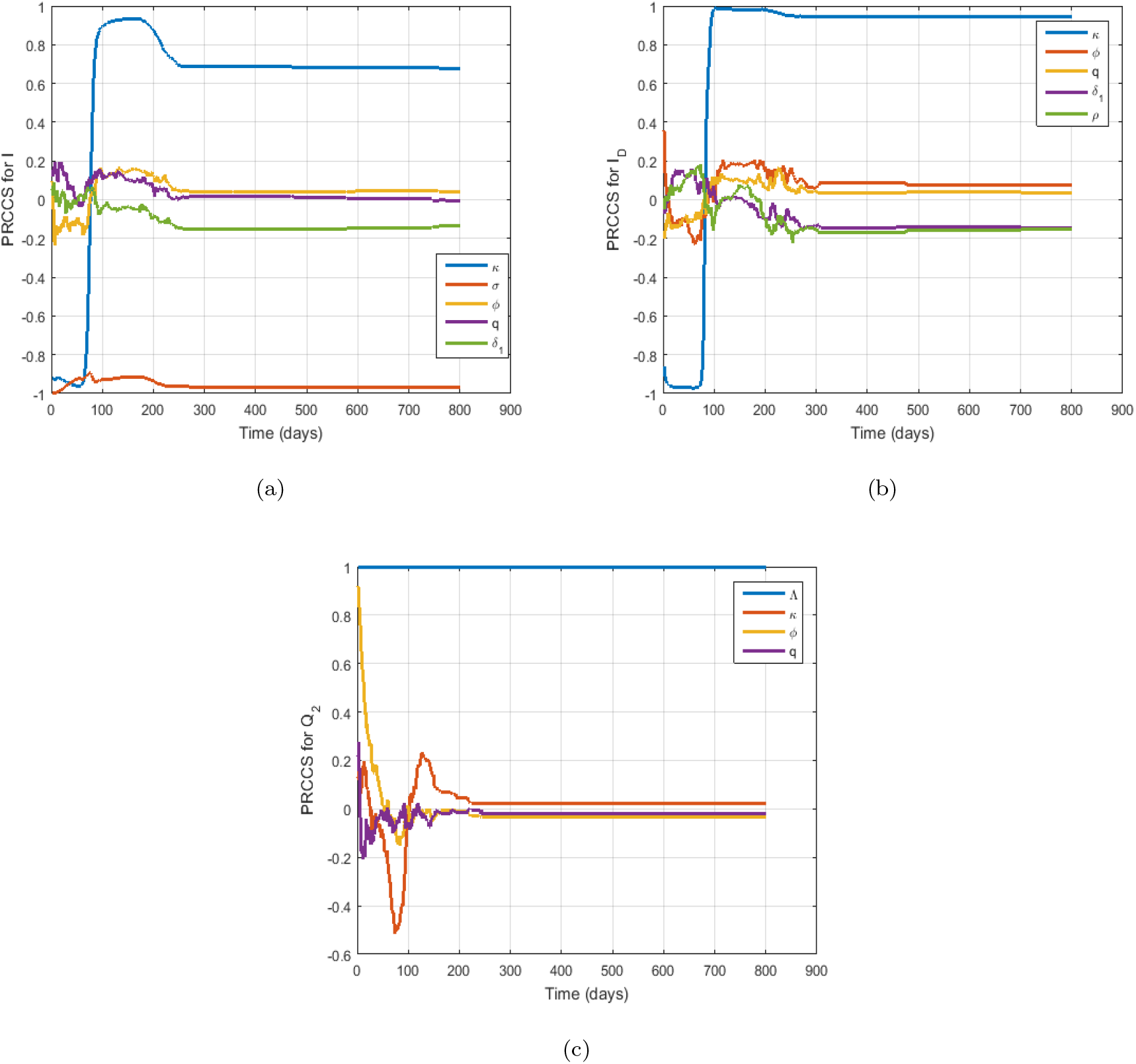
Plots of PRCC value showing the effects of long term dynamics before surge for parameter values on COVID-19 disease in Zimbabwe over time for (a) parameters *κ, σ, ϕ, q, δ*_1_ against *I*. (b) parameters *κ, ϕ, q, δ*_1_, *ρ* against *I*_*D*_. (c) parameters Λ, *κ, ϕ, q* against *Q*_2_.

##### Short term predictions dynamics

In Figure 7(a), we observe that the parameter, *κ* being the governmental role actions during the pandemic is negatively correlated at the onset of the disease and becomes positively correlated from *t* = 10 to *t* = 300 days. Meanwhile, the parameter, *σ* is negatively correlated over the modelling time with respect to the undetected individuals, *I*(*t*). Also, Figure 7(b) gives similar trend to what we observe in Figure 7(a), only that the parameter *ϕ* becomes positively correlated at the onset of the epidemic and diminishes as time progresses against the exposed individuals in quarantine, *I*_*D*_(*t*). On the other hand, we observe in Figure 7(c) that the immigration rate, Λ, has a perfect strong correlation, while the parameter, *ϕ*, is positive and diminishes with time. The parameter, *κ*, has a negative correlation from the time *t* = 0 to time *t* = 100 and a positive correlation from *t >* 100 and becomes less significant after *t >* 170.

##### Long term predictions dynamics

We observe in Figures 8(a) and 8(b) that at the onset of the disease, the parameter *κ* has a negative correlation and eventually becomes positive after time *t* = 80 for the undetected and exposed individuals in quarantine. Further, in Figure 8(c) *κ* is negatively correlated to the *Q*_2_ individuals and becomes positive after 90 days. The parameters, *σ* and Λ are strongly negative and positive correlated respectively against *I* and *Q*_2_ as observed in Figures 8(a) and 8(c) respectively. Biologically, this signifies that an increase in the immigration rate, Λ, will result to an increase in the number of quarantined individuals while a decrease in *σ* will lead to a decrease in the number of undetected individuals before a surge.

#### 4.2.2. Sensitivity analysis after surge

We also make use of Table 3 and a sample size of *N* = 500 runs with a unit step of 1 to perform sensitivity analysis under this subsection. Short term results are depicted in Figure 9 while long term results are depicted in Figure 10. The basic reproduction number for this case is found to be ℛ_0_ = 2.8.

**Figure 9:**
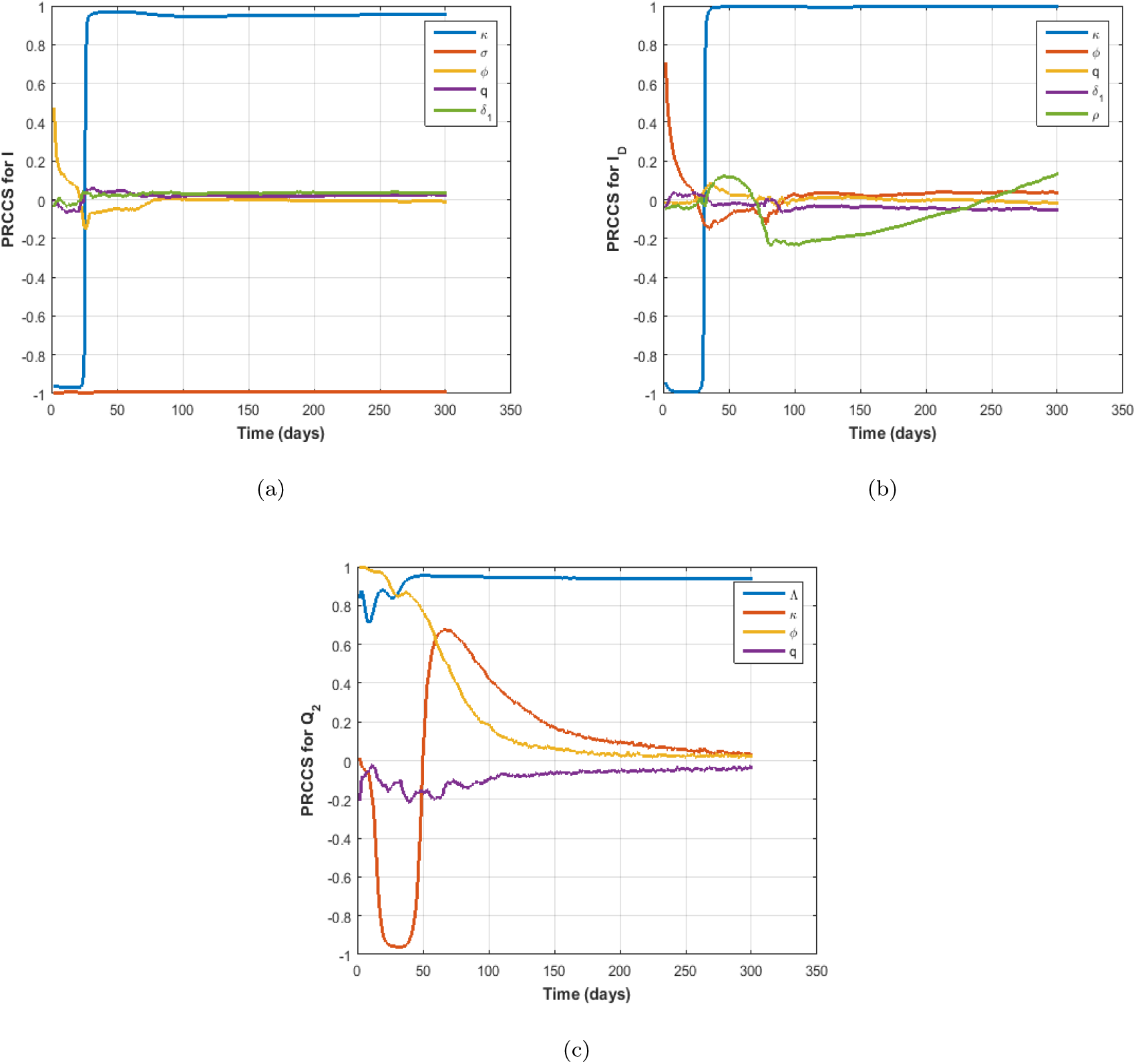
Plots of PRCC values showing the effects of short term dynamics after surge for parameter values on COVID-19 disease in Zimbabwe over time for (a) parameters *κ, σ, ϕ, q, δ*_1_ against *I*. (b) parameters *κ, ϕ, q, δ*_1_, *ρ* against *I*_*D*_. (c) parameters Λ, *κ, ϕ, q* against *Q*_2_.

**Figure 10:**
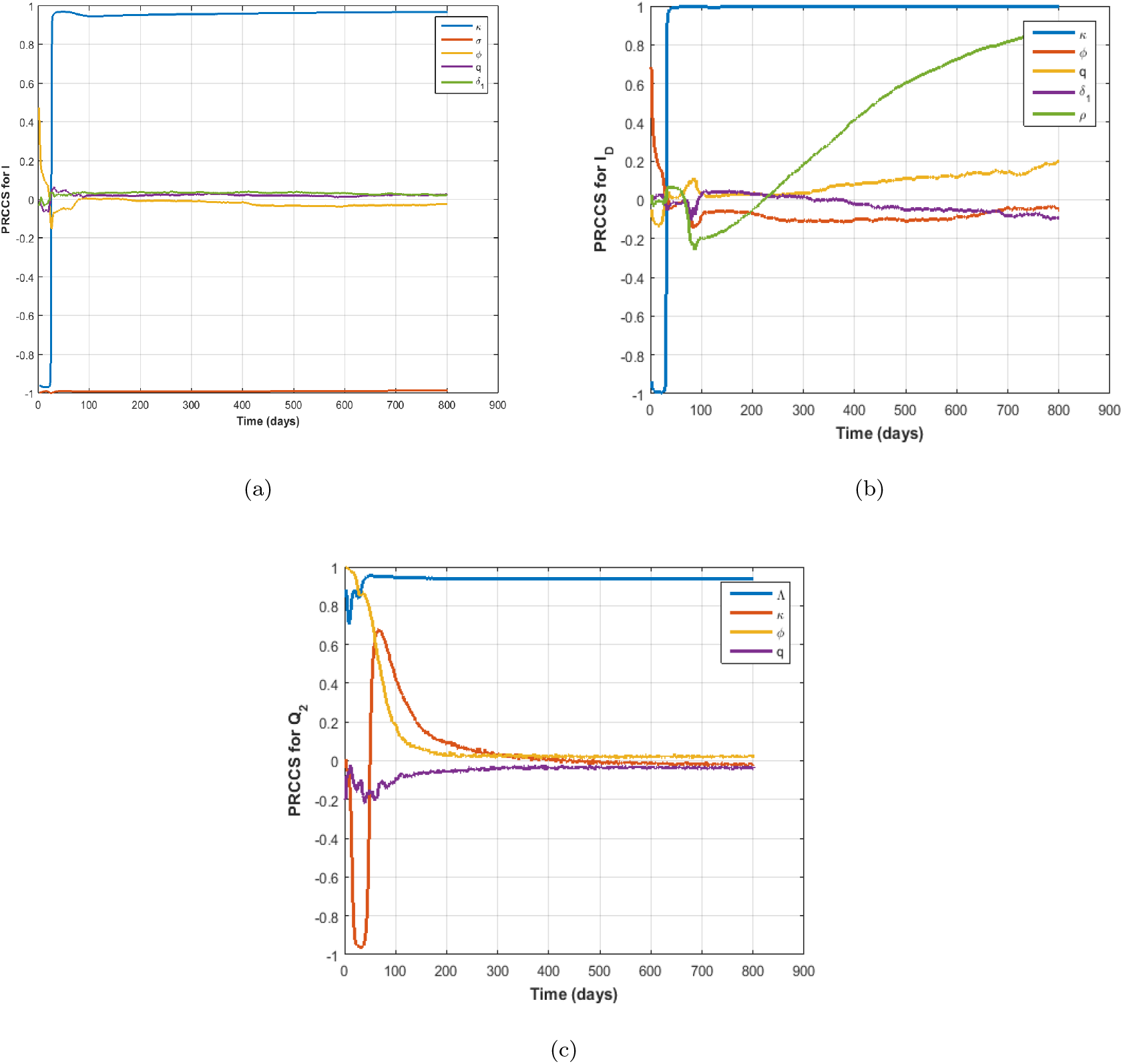
Plots of PRCC value showing the effects of long term dynamics after surge for parameter values on COVID-19 disease in Zimbabwe over time for (a) parameters *κ, σ, ϕ, q, δ*_1_ against *I*. (b) parameters *κ, ϕ, q, δ*_1_, *ρ* against *I*_*D*_. (c) parameters Λ, *κ, ϕ, q* against *Q*_2_.

##### Short term predictions dynamics

In Figures 9(a), 9(b) and 9(c), we observe that the parameter, *κ*, has a negative correlation from time *t* = 0 to *t* = 50 and thereafter it becomes positively correlated against the state variables *I, I*_*D*_ and *Q*_2_. This implies that an increase in governmental role action such as implementing lockdowns and wearing of mask needs to be continued after the first surge in order to control the disease effectively. On the other hand, the immigration rate, Λ, has a strong positive correlation over time as seen in Figure 9(c) while the parameter *σ* has a strong perfect negative correlation against the state variable, *I*_*D*_. Also, as seen before a surge, an increase in the immigration rate during COVID-19 will result in more infections in a wholly susceptible populations. On the other hand, the mean duration of quarantine parameter *ϕ* does not have a strong impact on the class, *I*_*D*_, as time increases as it becomes positive and diminishes.

##### Long term predictions dynamics

A similar trend is also seen in Figures 10(a) and 10(c) for the parameter *κ*. It has negative PRCC values from the first 30 days and positive PRCC values for *t >* 40. Also, in Figures 10(a), 10(b) and 10(c), we observe that the mean duration in quarantine, *ϕ*, has a positive PRCC value and diminishes as time progresses. The mean duration of incubation is not significant at the onset after the surge but become prominently positive correlated after time *t >* 200. In addition, the immigration rate, Λ, has a perfect positive correlation PRCC value over the modelling time.

### 4.3. Effects of varying parameters q and κ on I(*t*)

We investigate the impact of parameters *q* and *κ* on the number of undetected infected individuals, *I*(*t*). The impact of *q* is considered for both the period before and after the massive surge of returnees whereas the impact of *κ* is considered for the period after the massive surge of returnees.

We observe from Figure 11 that the variation of *q* before the massive surge of returnees has less impact as compared to the period after the massive surge of returnees. We note that an increase in the value of *q* fuels the presence of undetected infected individuals. A 20% increase in the value of *q* results in an increase of 10 more daily undetected infected cases who will be responsible for many undetected local transmissions. Figure 12 illustrates that an increase in the governmental action results in a decrease in the number of undetected infected individuals. We observe that a 20% increase in the intensity of *κ* will result in approximately a maximum drop of 15 daily undetected cases. This is a reflection of the importance of ensuring adherence to measures put in place by the government to help control this pandemic.

**Figure 11:**
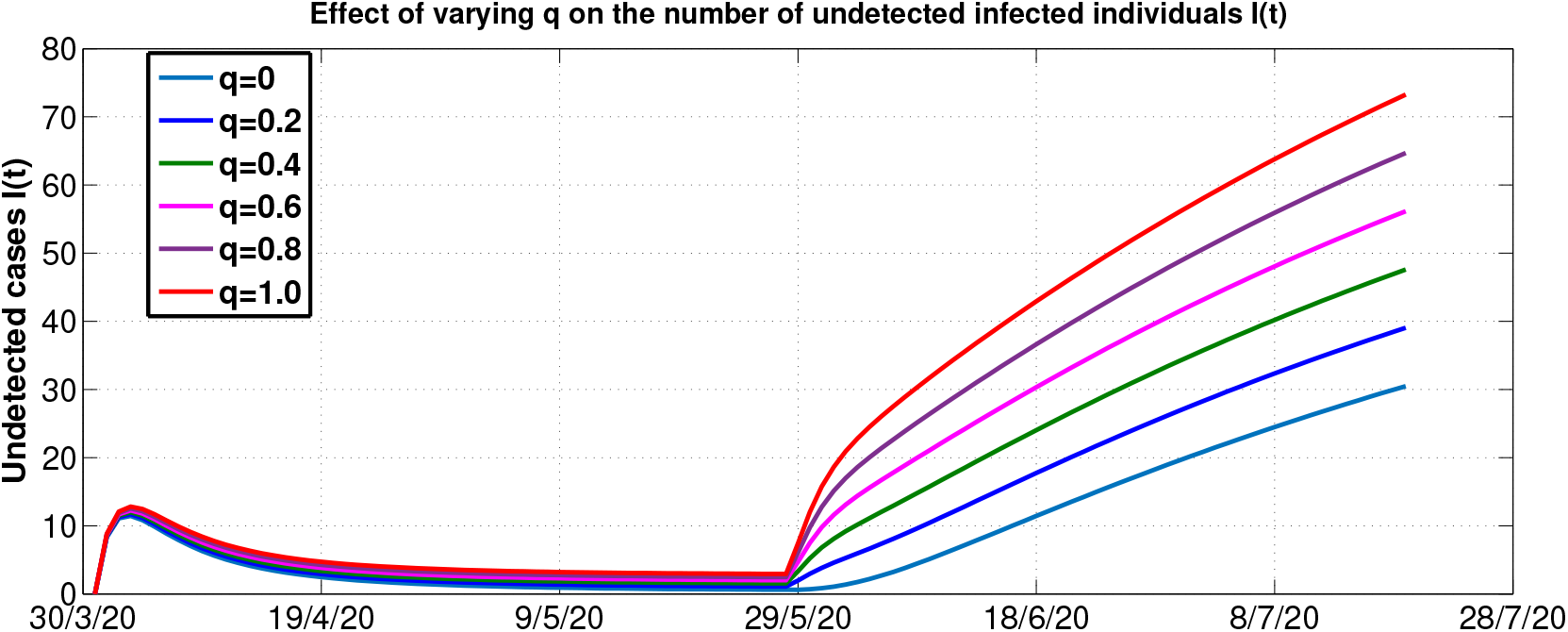
Effect of varying *q* on the number of undetected individuals.

**Figure 12:**
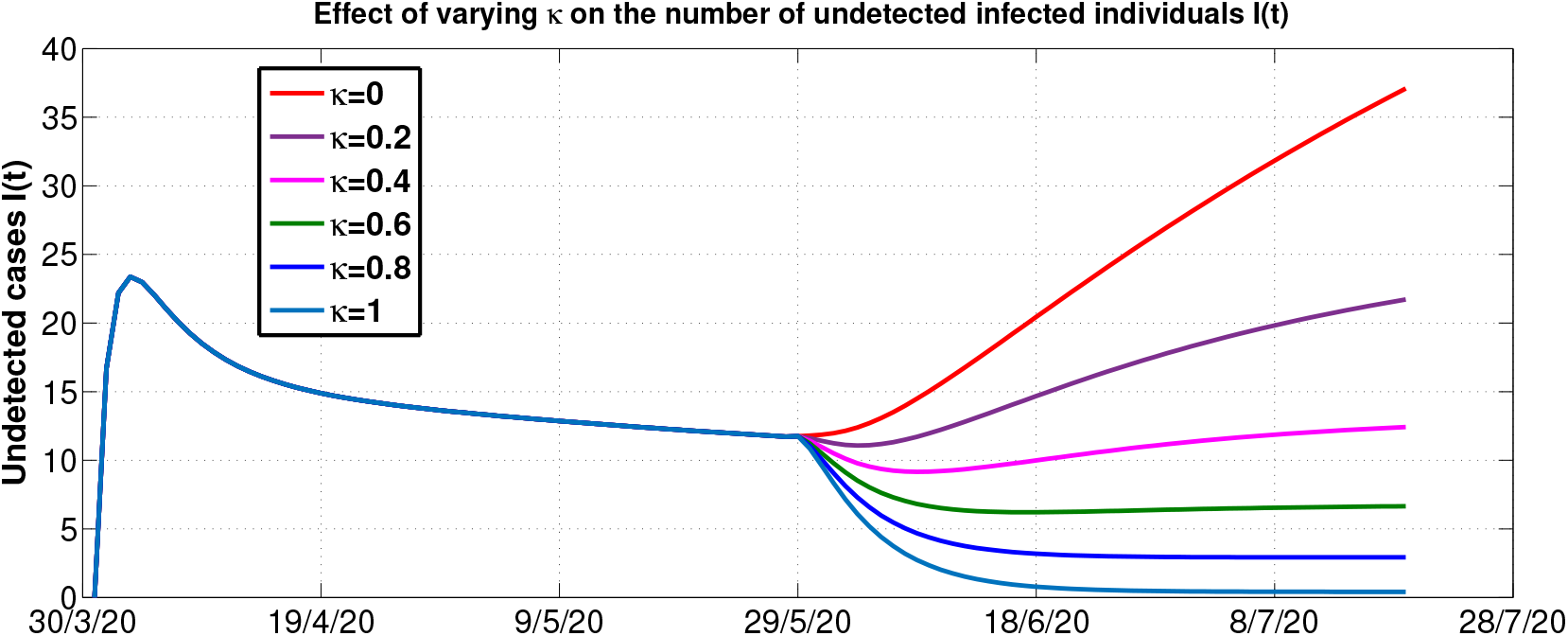
Effect of varying *k* after the 27th of May 2020 on the number of undetected individuals.

### 4.4. Effects of parameters on ℛ_0_

We use contour plots to investigate the effects of some crucial parameters on ℛ_0_. These results support the analytical findings of the sensitivity analysis done on ℛ_0_. Figures 13(a) and 13(c) show that an increase in the rate of implementation of governmental actions (*κ*) results in a decrease in the value of ℛ_0_. Also, we note that the instituted governmental policy of isolating detected infected individuals results in the reduction of ℛ_0_. Figure 13(b) shows that an increase in the proportion of escapees, *q*, will result in an increase in the disease transmission rate (*β*). This entails more infectious individuals as a result of escapees who will in turn increase local COVID-19 transmission.

**Figure 13:**
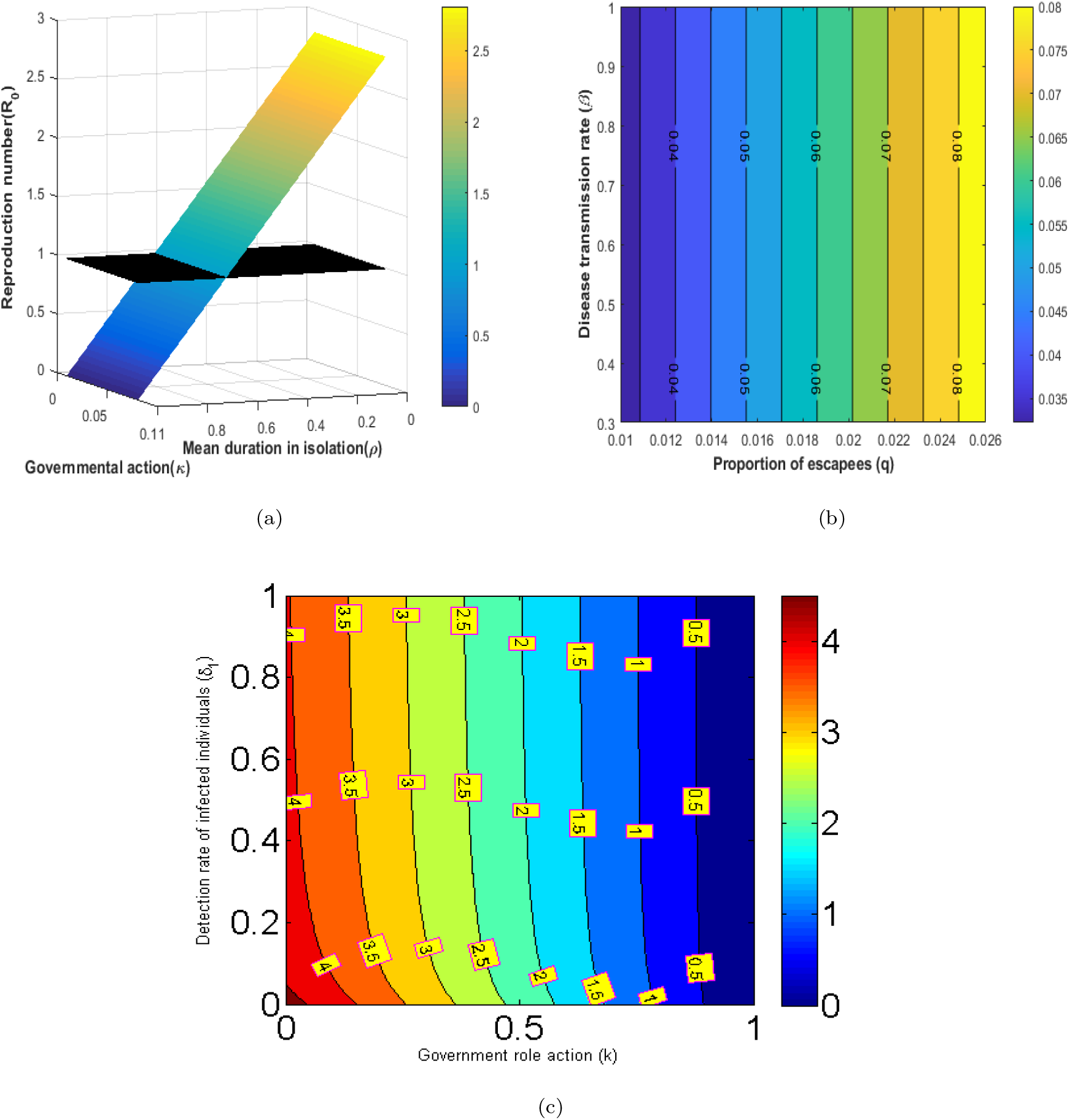
(a) 3D plot of parameters governmental action and mean duration in isolation versus the model basic reproduction number, ℛ_0_. (b) Contour plots of the model parameters proportion of escapees (*q*) versus disease transmission rate, *β*. (c) Contour plots of the model parameters governmental role action (*κ*) versus detection rate of infected individuals, *δ*_1_.

### 4.5. Effects of different lockdown measures

We investigate the impact of different lockdown scenarios on the COVID-19 dynamics in Zimbabwe. Figures 14, 15, 16 and 17 illustrate the impact of different lockdown strategies to curb the spread of the COVID-19 pandemic. Results show that the longer the period the lockdown is in force, the more reduction in the number of cases observed. However, this is usually accompanied by an economic impact that may be difficult to endure. Thus, some trade-offs between the economy and the infection would need to be taken care of, to ensure human life is preserved at a bearable economic cost. Besides the length of the lockdown period, we also note that the timing of the lockdown implementation is very crucial in the fight against the COVID-19 pandemic. A long delay in the implementation of lockdown can result in failure to minimize the arising number of infections. This also can lead to the number of infections increasing above the threshold healthcare capacity thereby resulting in an otherwise avoidable loss of life. Lastly, Figure 18 estimates the projected time for reaching peak cases of COVID-19 in the presence of the current dynamics of escapees. We observe that the peak number of cases will be reached by approximately the beginning of October 2020.

**Figure 14:**
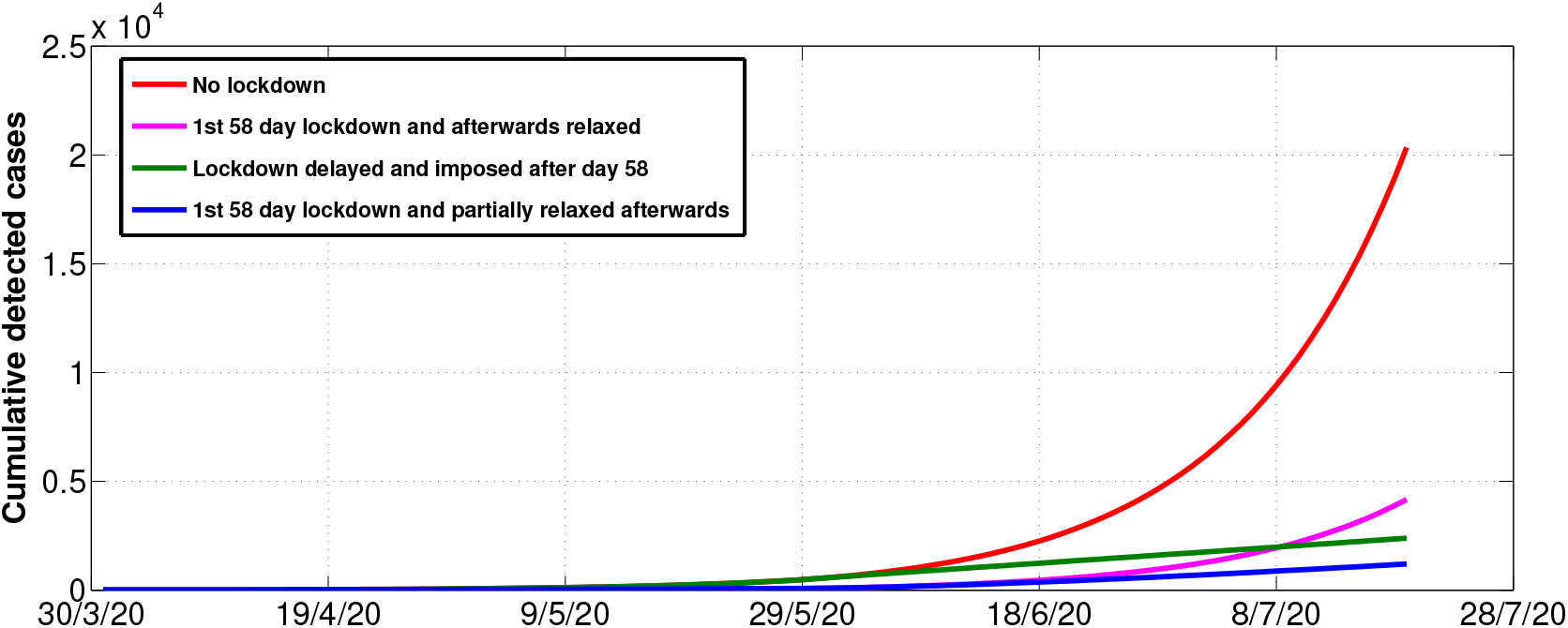
Simulation of cumulative detected cases under different lockdown scenarios.

**Figure 15:**
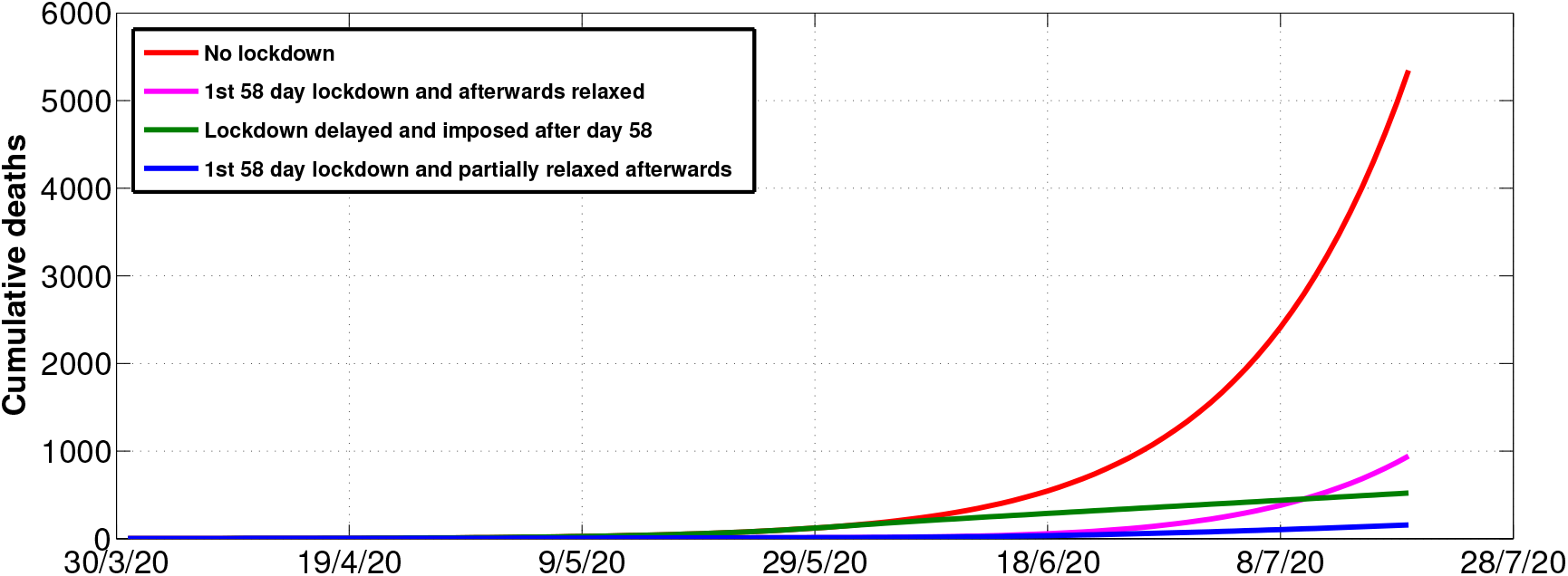
Simulation of cumulative deaths under different lockdown scenarios.

**Figure 16:**
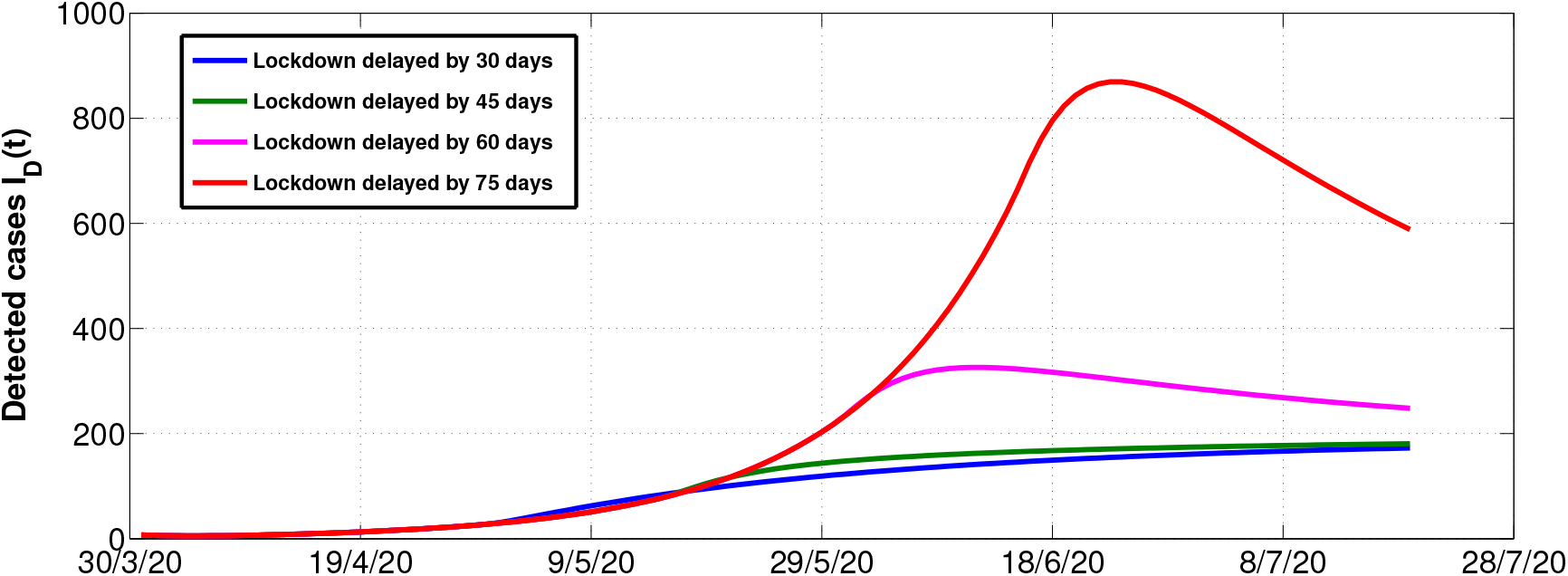
Simulation of daily detected cases for different delays in implementing lockdown measures.

**Figure 17:**
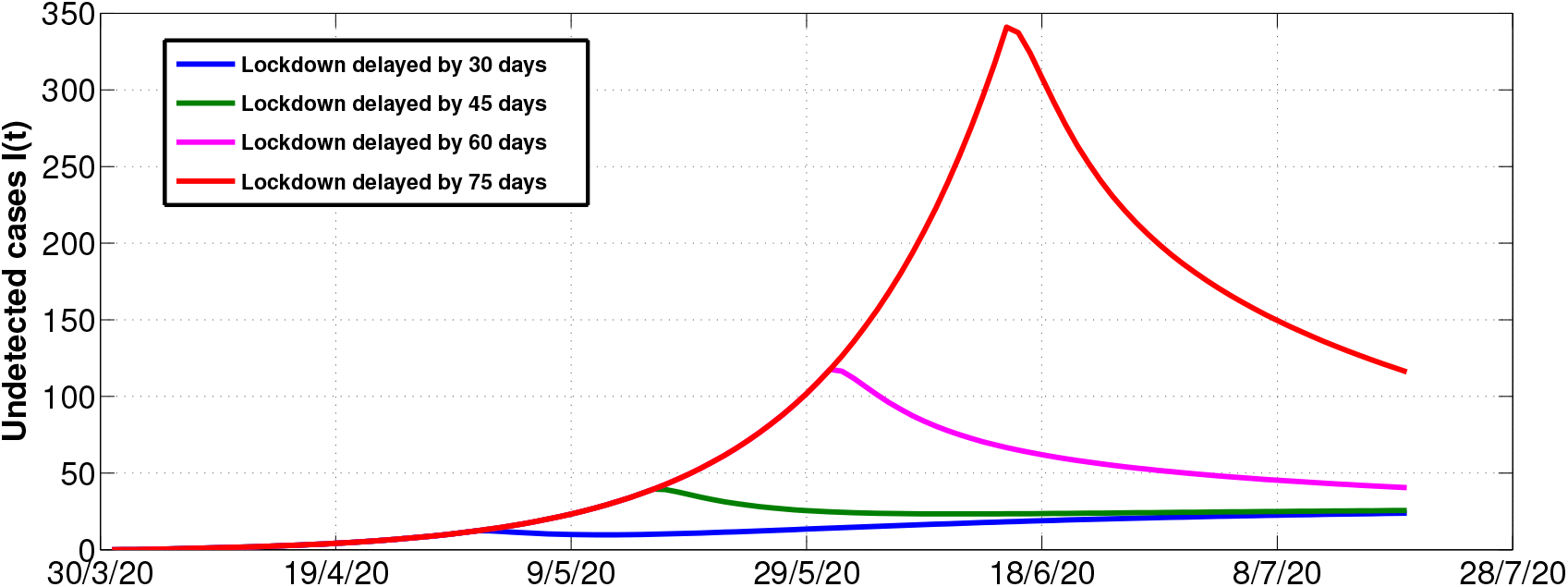
Simulation of daily undetected cases for different delays in implementing lockdown measures.

**Figure 18:**
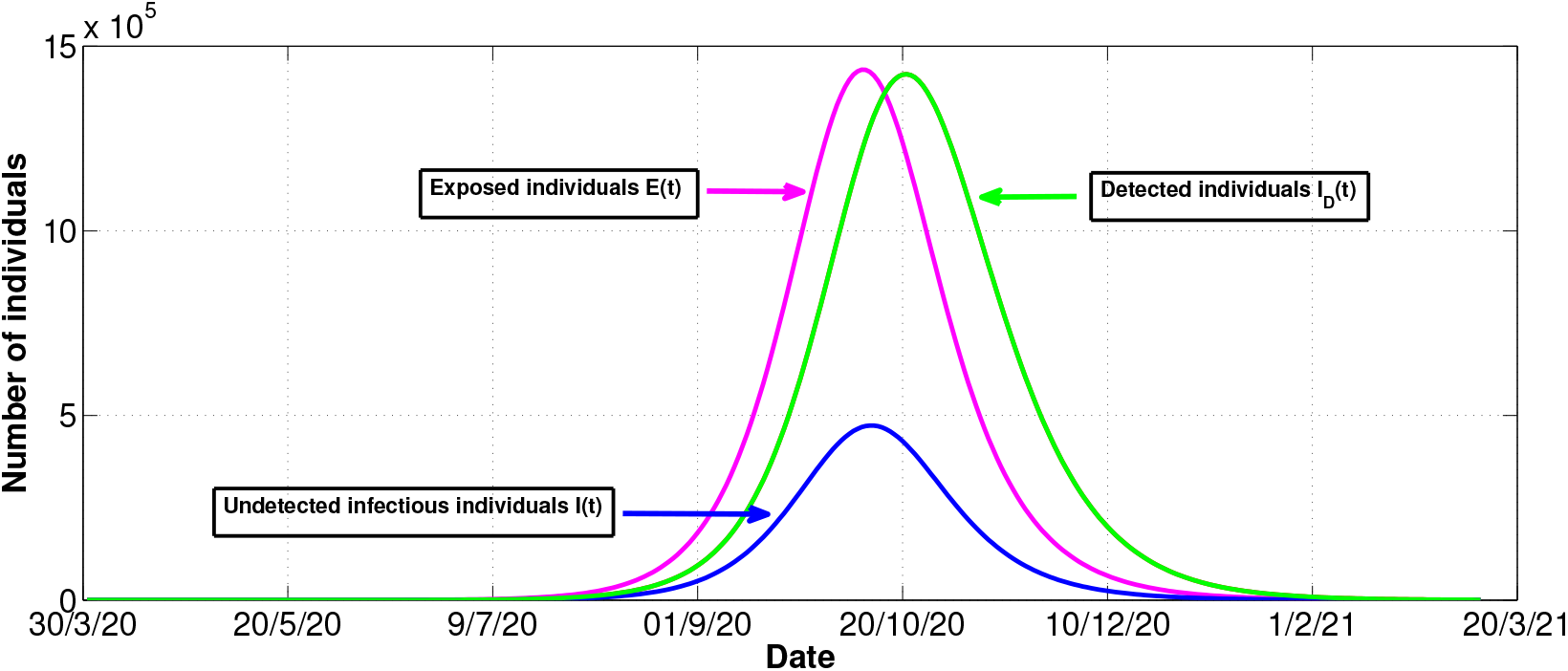
Simulation of projected peak cases of COVID-19 in the presence of escapees.

## 5. Conclusions

In this paper, we studied the role of escapees on the COVID-19 disease transmission dynamics in Zimbabwe using a deterministic compartmental model. The developed model is examined for important properties using tools drawn from the vast literature of mathematical epidemiology. The model reproduction number has been established and used to carry out stability analysis of the model.

Results from numerical simulations suggest that; first, an increase in the number of escapees from the quarantine centres will result in more local transmissions, which will lead to more people contracting the disease. Second, we found out that in order to contain the spread of the disease, governmental action of above 65% is required to curtail the spread of COVID-19 in Zimbabwe. Thus, it becomes paramount to implement measures that enhance adherence to intervention measures put in place by the government in order to curb this disease. Third, findings from the sensitivity analysis of the model indicate that the escapee parameter, *q* and the daily immigration rate, Λ, show a strong positive correlation to the number of daily reported new COVID-19 infections. This implies that a high number of escapees will result in more local transmissions and longer duration of the disease. Thus, mitigation measures such as increased surveillance in quarantine centres will be of great help in reducing the number of escapees.

The results presented in this manuscript are not without shortcomings. We did not consider the case of vital dynamics as the disease is about to reach its peak, hence incorporating vital dynamics will be very important. However, the results obtained in this research work are implementable and will influence policy management and decision making on the control of COVID-19 pandemic in Zimbabwe.

## Data Availability

Below website

https://reliefweb.int/report/zimbabwe/zimbabwe-situation-report-10-july-2020

## Acknowledgements

The authors acknowledge, with thanks, the support of their respective departments towards the production of this manuscript.

